# Urinary Schistosomiasis Among Primary School Children in Qua’an-Pan Local Government Area, Plateau State, Nigeria

**DOI:** 10.1101/2025.04.08.25325443

**Authors:** Waalmoep Gerald Damar, Morgak James Gonap, Gudzan Sow, Wandayi Emmanuel Amlabu

## Abstract

**Background:** Urinary schistosomiasis is a freshwater dependent urogenital infection that can be contracted by direct contact activities with water that contains the infective stage of *Schistosoma haematobium*. It is second only to malaria in its debilitating effects on humans and is a serious public health problem in Nigeria. Plateau State is classified as a hyper-endemic area.

**Objective:** This study was conducted to assess the prevalence of urinary schistosomiasis among primary school children in Qua’an-Pan Local Government Area, Plateau State.

**Method:** The cross-sectional study comprised of 806 children who provided midstream urine samples in clean labelled urine collection bottles. Sedimentation technique was used to prepare samples for parasitological examination. Data obtained was entered into SPSS version 20.0 for data analysis.

**Results:** The overall prevalence of urinary schistosomiasis in this study was 17.87%. Dokan Kasuwa district had the highest prevalence (27.5%) while the male children had a higher prevalence (20.29%) than the female children. Children between the ages of 15-19 years had a higher prevalence (24.53%) compared to the other age groups. Children attending private primary schools had a higher (18.51%) than their counterparts attending public primary schools (17.27%).

**Conclusion:** The prevalence of urinary schistosomiasis in Qua’an-Pan Local Government Area is lower than was previously reported but is still high, the situation should be closely monitored. More can be done to maintain the drop in prevalence by annual distribution of praziquantel tablets, provision of safe drinking water and creating awareness about the disease.

## Introduction

*Schistosoma haematobium* is a blood fluke and is the causative agent of urinary schistosomiasis among human beings^1^. Human urinary schistosomiasis is a water dependent urogenital infection that can be contracted from freshwater bodies that contain the parasite’s infected snail intermediate host *Bulinus* species^2^. Direct contact activities with water that contains the infective stage (cercariae) of the parasite aid the transmission of the disease to humans. Transmission will occur when the cercariae enters the skin of an individual to cause infection^3^. Blood in urine which in some cases is accompanied by inflammatory responses to the parasites eggs is a common symptom of the disease^4^. Other symptoms of the disease are dysuria, fibrosis of the bladder and urethra, hydronephrosis, haematemesis and irreversible lesions in the vulva, vagina, cervix and uterus which in turn can create an entry point for the human immune virus (HIV)^5,6^.

Urinary schistosomiasis was ranked second only to malaria in its debilitating effects on humans among the tropical diseases and it was third globally, after malaria and intestinal helminthiasis^7,8^. It is a serious public health problem in Nigeria with estimates showing that about 29 million people are infected and about 101 million people are at risk of infection with urinary schistosomiasis^9,10^. Recent estimates generated from the last 50 years classifies Plateau State as a hyper-endemic area (with prevalence predicted to be greater than or equal to 50%). Plateau State is ranked 12^th^ highest among the 36 states in Nigeria in terms of endemicity of the disease^8^. Dakul and colleagues^11^ reported a prevalence of 51% in Namu district of Qua’an-Pan Local Government Area (LGA). The study in Qua’an-Pan LGA was carried out over 20 years ago and there is a need to re-assess the prevalence of the infection in Qua’an-Pan LGA. This is needed because there have been efforts by Government and Non-Governmental organizations to treat and reduce the prevalence of the disease in Qua’an-Pan LGA. These efforts included the distribution of therapeutic drugs to school children (in four Nigerian states including Plateau State) by the Carter Center assisted programme, WHO, and Merck as well as the mapping of 17 states (Plateau State included) for schistosomiasis by the Federal Government of Nigeria^12,8^.

The aim of this study was to assess the prevalence of urinary schistosomiasis in Qua’an-Pan LGA, Plateau State, Nigeria. Findings from this study will provide information on the current status of urinary schistosomiasis in Qua’an-Pan LGA. The status of the infection will provide guidance and insight on the next steps to be taken in controlling the disease in Qua’an-Pan LGA.

## Method

### Study area

Qua’an-Pan LGA is made up of 8 districts and it is located in the southern part of Plateau State. It is located on latitude 8° 48□N and longitude 9° 09 □ E, covering an area of about 2,478 km^2^ and with an estimated population of 196,929 people^13^. It has a tropical climate and has an average annual temperature of 27.4 °C. Precipitation averages 1,235 mm and the average humidity is 52%. Temperatures vary by 4.5 °C throughout the year^14^.

### Study Design

The cross-sectional study was carried out from September, 2021 to December, 2021. A list of all the registered schools were obtained from the Education Board and Multi-stage random sampling was used to select children from five districts (Kwande, Kwalla, Dokan Kasuwa and Bwall Districts) in Qua’an-Pan LGA. Two schools (comprising a public and a privately-owned school) were selected from each district, making a total of 10 schools.

### Sample size

Sample size for the determination of urinary schistosomiasis in Qua’an-Pan LGA was calculated according Cochran’s formula^15^;

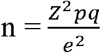

where;

n = sample size

z (standard normal distribution at 95% confidence interval) = 1.96

p (prevalence) = 51% ^11^

p = 0.51

q = 1 – p

q = 0.49

e (the allowable error, taken as 5%) = 0.05

The minimum sample size calculated for this study was 384 children, however, a total of 806 children participated in this study.

### Ethical consideration

The study was approved by the Committee on Use of Human Subjects for Research, Ahmadu Bello University, Zaria (ABUCUHSR/2021/030). Informed signed consent letters were obtained from the parents/guardians of the selected children. Also, informed oral consents were obtained from the Head-teachers and children that participated in this study. Children whose parents didn’t sign consent forms were not allowed to participate in the study. The children were informed of their right to withdraw from the study at any time without consequence.

### Sample collection

Each participant was given a clean labelled urine collection bottle and given instructions on how to transfer mid-stream urine into the bottle. About 2 ml of formalin was added to each urine sample for fixation of the parasite eggs^16^. The urine samples were kept in a box and transported to the Helminthology Laboratory of the Faculty of Veterinary Medicine (Ahmadu Bello University, Zaria, Nigeria) for parasitological examination. Samples were kept in a refrigerator before they are analyzed. The age and sex of each child were documented on submission of their urine samples.

### Examination of Urine Samples

The urine samples were examined using the sedimentation method^6^. About 10–30 ml of urine samples received from the children was thoroughly mixed after which a 10 ml aliquot was transferred into a centrifuge tube and it was spun at 5,000 rpm for 5 minutes. The supernatant was decanted, while a drop of the sediment was placed on a clean grease free slide and covered with a cover slip. The urine samples on the glass slides were examined under the microscope using the 10× and 40× objectives. Identification of the *Schistosoma haematobium* eggs was done using the WHO (1994) Bench Aid^17^.

### Data analysis

Prevalence was calculated using percentages while Chi-square test was used to determine significant difference between variables and p ≤ 0.05 was considered statistically significant. The SPSS version 20.00 was used to analyze data.

## Results

### Prevalence of urinary schistosomiasis based on districts in Qua’an-Pan Local Government Area

Table 1 presents the prevalence of urinary schistosomiasis across the five districts that were sampled during the study. The overall prevalence of urinary schistosomiasis among primary school children in Qua’an-Pan LGA was 17.87%. Dokan Kasuwa district had the highest prevalence (23.13%) of urinary schistosomiasis while the lowest prevalence (14.29%) was observed in Kwande District. There was no statistical significance in the prevalence among the districts (χ^2^= 6.11, p = 0.19).

**Table 1:** Prevalence of urinary schistosomiasis based on districts among children in Qua’an-Pan Local Government Area.

| District | Number examined | Number negative | Number positive | % positive |
| --- | --- | --- | --- | --- |
| Kwalla | 159 | 135 | 24 | 15.09 |
| Bwall | 169 | 135 | 34 | 20.12 |
| Kwang | 136 | 113 | 23 | 16.91 |
| D.K. | 160 | 123 | 37 | 23.13 |
| Kwande | 182 | 156 | 26 | 14.29 |
| Total | 806 | 662 | 144 | 17.87 |

Key: D.K.= Dokan Kasuwa. $\chi^2 = 6.11$ ; $df = 4$ ; $p = 0.19$ .

### Prevalence of urinary schistosomiasis based on sex among primary school children in Qua’an-Pan Local Government Area

Table 2 presents the results of urinary schistosomiasis among males and females. The table shows that 20.29% of males and 15.36% of females were positive for urinary schistosomiasis. There was no statistically significant difference in infection between males and females (χ^2^ = 3.33; p = 0.08).

**Table 2:**
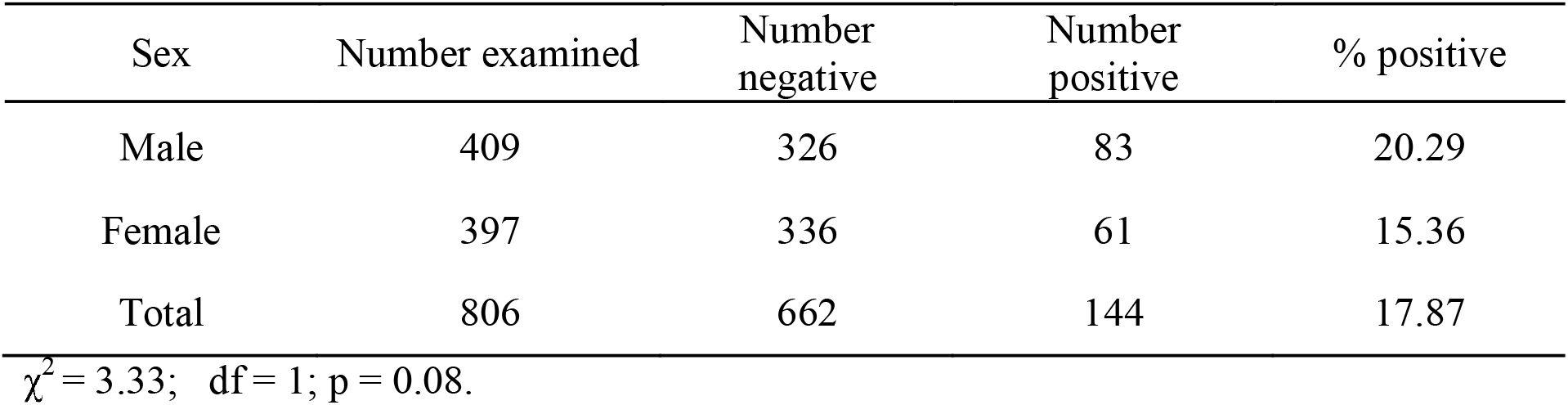
Prevalence of urinary schistosomiasis based on sex among children in Qua’an-Pan Local Government Area.

### Prevalence of urinary schistosomiasis among children based on age group in Qua’an-Pan Local Government Area

The prevalence of urinary schistosomiasis among the three age groups (5-9, 10-14 and 15-19 years) is presented in Table 3. When the data was stratified by age group, the highest prevalence was observed in the 15-19 years age group (24.53%) and the lowest prevalence was observed in the 10-14 years age group (16.67%). However, the differences in prevalence among age groups were not statistically significant (χ^2^ = 2.20, p = 0.33).

**Table 3:** Prevalence of urinary schistosomiasis based on age group among children in Qua’an-Pan Local Government Area.

| Age group (years) | Number examined | Number negative | Number positive | % positive |
| --- | --- | --- | --- | --- |
| 5 – 9 | 273 | 222 | 51 | 18.68 |
| 10 – 14 | 480 | 400 | 80 | 16.67 |
| 15 – 19 | 53 | 40 | 13 | 24.53 |
| Total | 806 | 662 | 144 | 17.87 |

$\chi^2 = 2.20$ ; $df = 2$ ; $p = 0.33$ .

### Prevalence of urinary schistosomiasis among children attending public and private primary schools in Qua’an-Pan Local Government Area

Table 4 presents the prevalence of urinary schistosomiasis among children attending two types of primary schools: the public and private owned schools. The results show that children attending private schools had a slightly higher prevalence than children in public schools, with no significant difference observed between children in public and private schools (χ^2^ = 0.21, p = 0.65).

**Table 4:** Prevalence of urinary schistosomiasis among children attending public and private primary schools in Qua’an-Pan Local Government Area.

| School | Number examined | Number negative | Number positive | % positive |
| --- | --- | --- | --- | --- |
| Public | 417 | 345 | 72 | 17.27 |
| Private | 389 | 317 | 72 | 18.51 |
| Total | 806 | 662 | 144 | 17.87 |

$\chi^2 = 0.21$ ; $df = 1$ ; $p = 0.65$ .

## Discussion

The overall prevalence of urinary schistosomiasis in this study was 17.87%. The prevalence recorded in this study falls within the range where it is recommended that about 75% of the population receive annual preventive chemotherapy with single dose praziquantel tablets^18^. It is lower than the previous prevalence reported in a longitudinal study carried out in Namu district of Qua’an-Pan LGA that presented prevalence of 65.8% and 51.5% in 1995 and 1996 respectively^11^. The prevalence recorded in this study is higher than the prevalence reported in Jos North LGA, Mangu LGA and Ibadan in Oyo State, Nigeria^19,20,21,22^. These locations can be classified as semi-urban or urban settings with access to pipe borne water and people in such locations depend less on streams and rivers for water. It will not be out of place to state that the overall prevalence recorded in this study is lower than what was estimated. This is because Plateau State was classified as a hyper-endemic region. The low prevalence could be as a result of irregular distribution of praziquantel tablets to schools with special attention given to hyper-endemic areas such as Plateau State^8^. The low prevalence could also be an indication of the increasing level of awareness about the disease in the study area. It could also be because the cross-sectional study was carried out during the end of the rainy season and the beginning of the dry season. During this period, severe water scarcity has not yet set in and there is less dependence on rivers and streams for water as the wells in the homes still have water in them.

The male children had a higher but statistically insignificant prevalence of urinary schistosomiasis than the female children. This disagrees with the work of Banwat *et al*.^21^ in Mangu LGA of Plateau State who reported a significantly higher prevalence in male children. The similar prevalence observed between both sexes be because both male and female children take part in similar water contact activities such a swimming, bathing, washing and farming.

The highest prevalence of urinary schistosomiasis was observed in the 15-19 years age group. A similar finding of older children having a higher prevalence than younger children was also observed in Benue State^23^. The higher prevalence observed among the older children could be because farmers tend to employ the services of the older children on their farm. One of the major crops grown in the study area is rice and it requires a swampy area to grow well. While farming, the children spend long periods in direct contact with water and it increases the risk of infection with the causative parasite.

The children that attended private primary schools had a slightly higher but statistically insignificant prevalence of urinary schistosomiasis than their counterparts in the public primary schools. This observation implies that children irrespective of the type of school they attended in the study area were equally exposed to the infection. This also suggests that the children in both types of schools took part in similar activities or exhibited similar behaviours in relation to water contact activities.

## Conclusion

Qua’an-Pan LGA is endemic for urinary schistosomiasis despite the reduction in prevalence reported in this study. The reduction in prevalence offers encouragement in the fight against the disease but there is need to maintain the annual distribution of praziquantel tablets in schools. Also, the results show that sex, age group, type of school and location were not risk factors for the disease.

## Data Availability

All data produced in the present study are available upon reasonable request to tye authors

## Limitations

This study was not able to investigate socio-economic traits and behavioural factors that could serve as risk factors for urinary schistosomiasis in the study area. A study encompassing these factors can provide information that will guide policies and interventions against the menace of urinary schistosomiasis.

## Acknowledgements

We would like to highlight the work of God the ultimate author. The children, teachers, Head-teachers and parents/guardians that participated in this study were our heroes. This work is dedicated to them for their input, understanding and cooperation. Special appreciation goes to Mr. Wilberforce Sogotdiel who took the time to be our guide and help with sample collection.

